# Acute to long-term characteristics of impedance recordings during neurostimulation in humans

**DOI:** 10.1101/2024.01.23.24301672

**Authors:** Jie Cui, Filip Mivalt, Vladimir Sladky, Jiwon Kim, Thomas J. Richner, Brian N. Lundstrom, Jamie J. Van Gompel, Hai-long Wang, Kai J. Miller, Nicholas Gregg, Long Jun Wu, Timothy Denison, Bailey Winter, Benjamin H. Brinkmann, Vaclav Kremen, Gregory A. Worrell

**Author notes:** ORCID: 0000-0003-1000-8869), ORCID: 0000-0002-0693-9495), ORCID: 0000-0002-4712-7039), ORCID: 0000-0002-5310-5549), ORCID: 0000-0001-8087-7870), ORCID: 0000-0001-9013-3007), ORCID: 0000-0002-6151-043X), ORCID: 0000-0001-8019-3380), ORCID:0000-0002-5404-4004), ORCID: 0000-0003-1157-2138), ORCID: 0000-0002-2392-8608), ORCID: 0000-0001-9844-7617), ORCID: 0000-0003-2916-0553). Corresponding Author: Gregory A. Worrell MD PhD, Professor of Neurology and Biomedical Engineering Consultant, Neurology Mayo Clinic, Rochester, MN 55905.

## Abstract

**Objective:** This study aims to characterize the time course of impedance, a crucial electrophysiological property of brain tissue, in the human thalamus (THL), amygdala-hippocampus (AMG-HPC), and posterior hippocampus (post-HPC) over an extended period.

**Approach:** Impedance was periodically sampled every 5-15 minutes over several months in five subjects with drug-resistant epilepsy using an experimental neuromodulation device. Initially, we employed descriptive piecewise and continuous mathematical models to characterize the impedance response for approximately three weeks post-electrode implantation. We then explored the temporal dynamics of impedance during periods when electrical stimulation was temporarily halted, observing a monotonic increase (rebound) in impedance before it stabilized at a higher value. Lastly, we assessed the stability of amplitude and phase over the 24-hour impedance cycle throughout the multi-month recording.

**Main results:** Immediately post-implantation, the impedance decreased, reaching a minimum value in all brain regions within approximately two days, and then increased monotonically over about 14 days to a stable value. The models accounted for the variance in short-term impedance changes. Notably, the minimum impedance of the THL in the most epileptogenic hemisphere was significantly lower than in other regions. During the gaps in electrical stimulation, the impedance rebound decreased over time and stabilized around 200 days post-implant, likely indicative of the foreign body response and fibrous tissue encapsulation around the electrodes. The amplitude and phase of the 24-hour impedance oscillation remained stable throughout the multi-month recording, with circadian variation in impedance dominating the long-term measures.

**Significance:** Our findings illustrate the complex temporal dynamics of impedance in implanted electrodes and the impact of electrical stimulation. We discuss these dynamics in the context of the known biological foreign body response of the brain to implanted electrodes. The data suggest that the temporal dynamics of impedance are dependent on the anatomical location and tissue epileptogenicity. These insights may offer additional guidance for the delivery of therapeutic stimulation at various time points post-implantation for neuromodulation therapy.

## Introduction

Implantable neural sensing and stimulation (INSS) devices, which are capable of closed-loop therapy based on continuous monitoring of brain local field potentials and automated brain state classifications, enable adaptive long-term neural modulation [1–5]. Thus, the stability of the electrode-tissue interface is needed for optimal maintenance of accurate automated brain-state classifications, such as sleep and seizure, and optimal neuromodulation therapy over long periods (months to years) [6–8]. Instability of the electrode-tissue interface and associated electrical impedance may compromise the quality of electrophysiological recordings, brain-state classification, and the delivery of appropriate therapy.

Electrode impedance is widely used to assess the quality and stability of neural sensing and stimulation [9]. Changes in electrical interface impedance are generally assumed to stabilize multiple weeks after implantation [10, 11]. Impedance determines the local field potential (LFP) characteristics [12], which have been used in automated brain-state classifications, and drives the voltage-current relationship of therapeutic electrical stimulation. Impedance measurements can be conveniently performed using sensing and stimulation electrodes. Impedance changes provide insights into time-varying factors related to devices and tissues but may indicate problems with the electrode or electrode-tissue interface [7]. Although short-term changes in electrical impedance post-implant are well documented [7, 13], reports of long-term impedance measurements in humans are sparse [6] and relatively little is known about the chronic characteristics of impedance in the human limbic system [14, 15]. However, this information is necessary for evaluation and optimal tracking of biomarkers and delivery of therapeutic stimulation [2].

In addition to its role in interrogating INSS, electrical impedance is useful for characterizing brain neurophysiology [15]. For instance, in the central nervous system (CNS), increased impedance is correlated with decreased extracellular space (ECS) [16–18], modified electric field potential propagation [12], increased seizure activities [19, 20], and behavioral state changes [8, 18, 21–23].

In this study, we analyzed long-term impedance recordings from five patients with drug-resistant epilepsy implanted with investigational INSS devices [2]. The electrical impedance was measured periodically, as described in the Methods section. We investigated acute (1 – 3 days), subacute (4 days to 3 weeks post-implant), and long-term (> 3 weeks) two-point impedance measurements from multiple brain regions. We compared a piecewise and continuous mathematical model to capture the initial drop and later recovery phases of acute and subacute impedance dynamics.

We analyzed impedance changes during *gaps* in therapeutic stimulation. Previous studies have indicated that repeated voltage biasing can reduce electrode-tissue impedance [24]. Therefore, measuring impedance change during brief pauses in therapeutic electrical stimulation can provide insights into impedance change at electrode-tissue interface.

Finally, we described long-term amplitude and phase of 24-hour^1^ cycle of impedance [23]. Spectral analysis of impedance timeseries revealed strong oscillations around the periodicity of 24.01 ± 0.39 hour, consistent with previously observed extracellular volume changes in rodent glymphatic system [18]. We thus analyzed the long-term characteristics of the impedance cycle as indicators of their stability.

## Methods

### Subjects and data acquisition

Subject recruitment, INSS implantation, and electrical impedance measurements were performed as previously described [2, 23]. Briefly, five human subjects (S1 – S5) with drug-resistant bilateral mesial temporal lobe epilepsy were implanted with investigational Medtronic Summit RC+S™ devices (Medtronic, Minnesota, USA) targeting the bilateral anterior nucleus of the thalamus and bilateral mesial temporal structures. While the patients had bilateral mesial temporal lobe epilepsy, it is important to note that the left hemisphere was the most epileptogenic, with most seizures and interictal epileptiform activity (Table 1). Each patient was implanted with four leads with four electrodes (contacts) per lead (16 channels per patient); leads were implanted in the left and right thalamus (THL), amygdala-hippocampus (AMG-HPC), and posterior hippocampus (post-HPC), except for S2, whose left AMG-HPC was partially resected from a prior anterior temporal lobectomy.

**Table 1.** Summary of impedance recording and types of seizures.

| Subject | Days | Samples/Day | Sampling<br>interval (min) | Left<br>Sz | Right<br>Sz | Both<br>Sides Sz | Self-Rep.<br>Sz | Total<br>Number |
| --- | --- | --- | --- | --- | --- | --- | --- | --- |
| S1 | 693.49 | 57.44 | 25.05 ± 126.45 | 537 | 1 | 5 | 1 | 544 |
| S2 | 284.99 | 33.13 | 43.37 ± 289.76 | 3728 | 11 | 2 | 17 | 3758 |
| S3 | 182.28 | 74.46 | 19.28 ± 32.53 | 66 | 45 | 1 | 0 | 112 |
| S4 | 294.57 | 74.74 | 18.76 ± 84.97 | 39 | 0 | 0 | 0 | 39 |
| S5 | 124.06 | 239.88 | 5.97 ± 45.37 | 129 | 19 | 1 | 0 | 149 |

Abbreviation: Sz, Seizure; Self-Rep., self-reported

Platinum-Iridium (Pt-Ir) alloy contacts were used, owing to their low impedance, electrochemical stability, excellent biocompatibility, corrosion resistance, and radiopacity. They are widely used as implanted pial cortical and parenchymal electrodes for electrical brain stimulation (continuous, duty cycle, and responsive stimulation) with charge-balanced, asymmetrical biphasic Lily pulse waveforms over a wide range of frequencies (∼2 – 145 Hz) within established safe charge densities (< 30 µC/cm2). Two types of leads were implanted, with Medtronic 3387 leads (span 10.5 mm with four 1.5 mm long and 1.27 mm diameter contacts, surface area = 5.985 mm^2^, separated by 1.5 mm) targeting bilateral THL areas and Medtronic 3391 leads (spans 14.5 mm with four 3 mm long and 1.27 mm diameter contacts, surface area = 11.97 mm^2^, separated by 4 mm) targeting bilateral AMG-HPC and post-HPC areas. A longer span of Medtronic 3391 lead was used along the long axis of the AMG-HPC complex.

Electrical two-point monopolar measurements were nonuniformly sampled in these five subjects and streamed to the cloud database through a wireless network [2, 25]. The impedance measurement methods are detailed previously [23] and a brief description is provided in Figure 1. The monopolar impedances were sampled from the 16 electrode contacts using the RC+S™ device as the monopolar current return for using a single square-wave current pulse (0.4 mA, 80 µs pulse width). The voltage (*V*) was measured at 70 µs near the end of the injected pulse (*I*), and the effective impedance (Z) was calculated as *Z* = *V*/*I* following Ohm’s law. Our benchtop experiments show that this effective impedance is equivalent to an impedance measured by injecting a 1 kHz sinusoidal current with an amplitude of 500 nA, insensitive to the electrode-tissue (or electrode-electrolyte) interface impedance. Current stimulation and sensing voltage values were delivered using the same electrodes, while therapeutic stimulation was delivered only via 3387 leads targeting the THL (two per hemisphere).

**Figure 1.**
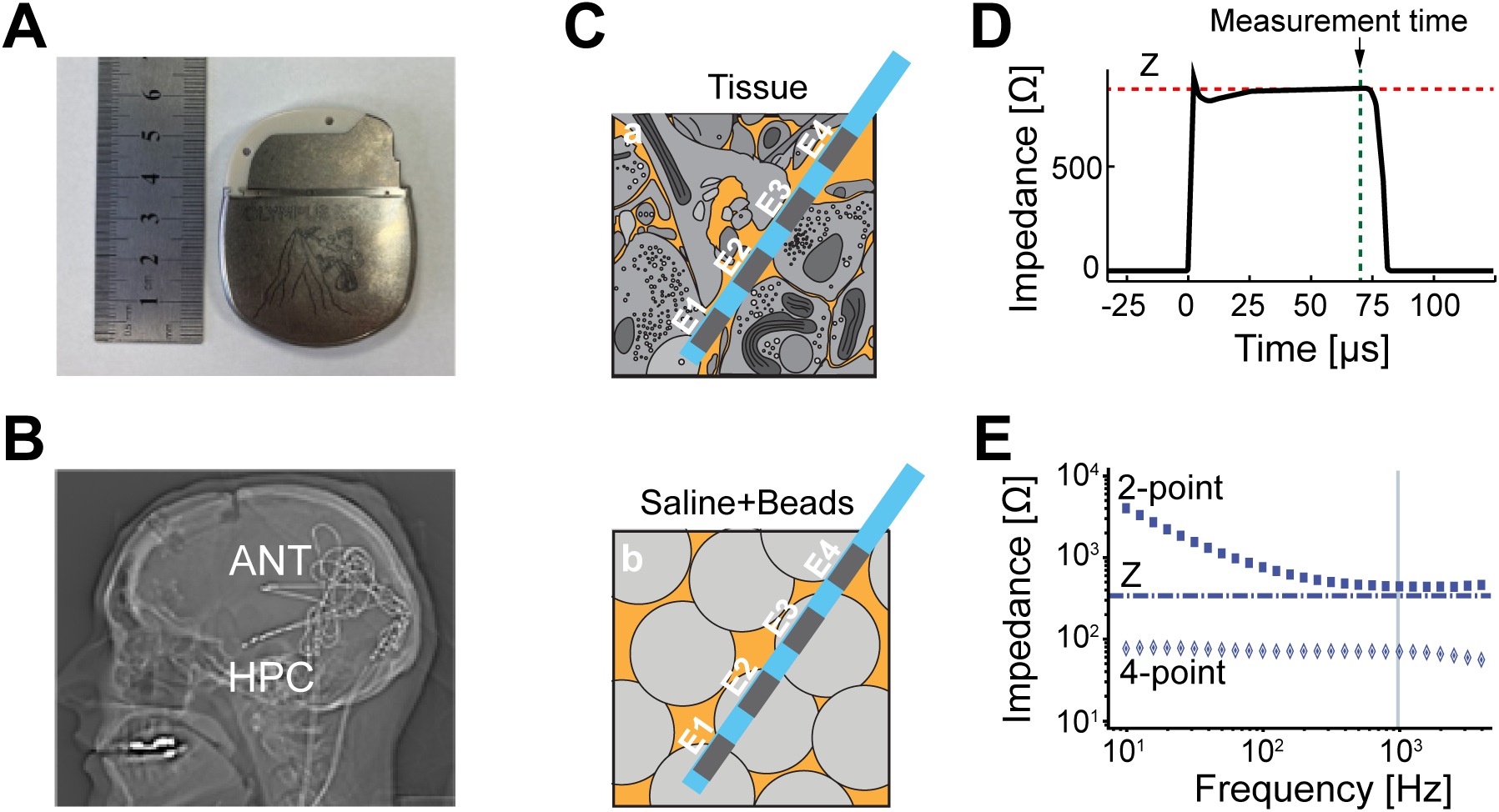
Impedance measurement. **(A)** The investigational Medtronics RC+S™ is a rechargeable device that enables 16 electrode contact electrical stimulation and programmable 4 LFP sensing channels from bipolar electrode contact pairs. **(B)** Lateral x-ray after implantation of the bilateral ANT (3387-leads) and bilateral AMG-HPC (3391-lead) targets. The lead extensions are tunneled down the neck to the sub-clavicular device pocket. The 3391-lead has four contacts (surface area = 11.97 mm2) spanning 24.5 mm. The contacts are 3.0 mm long and separated by 4.0 mm. The 3387-lead has four contacts (contact surface area = 5.985 mm2) spanning 10.5 mm. The individual contacts are 1.5 mm long and separated by 1.5 mm. **(C)** Schematic diagrams of the microenvironment of the electrode and brain tissue (a) and the corresponding model using saline/microbead composites (b) for benchtop experiment. The 2-point measurement employs the same electrodes contacts (E1 & E2) for both electrical stimulation and voltage sensing. The 4-point impedance measurement uses different electrodes for stimulation (E1 & E4) and sensing (E2 & E3). The 4-point measurements eliminate the interface electrode-electrolyte polarization, related to electrical stimulation, from the voltage measurements. **(D)** The RC+S™ calculates 2-point impedance using Ohm’s law, Z = V/I, where *I* is the injected current (0.4 mA, 80 µs pulse width) and *V* is the voltage response measured at 70 µsec. The voltage recording using 2-point measurement shows the voltage response to the impulse current (0.4 mA, 80 µs pulse width) with charging of the electrode-electrolyte double layer capacitor, which reaches an asymptotic voltage within ∼50 µs. **(E)** Impedance measured using sinusoidal currents in saline/microbead composites (1 – 5000 Hz). The 2-point measurements are dominated at low frequency (< 500 Hz) by the frequency dependent capacitive double-layer related to the electrolyte polarization at the electrode-electrolyte interface. The 4-point impedance measurement, utilizing different electrodes for current injection and voltage response sensing, yields a purely resistive impedance with no frequency dependence (10 – 5000 Hz). The RC+S™ impedance measurement (blue dashed line) can be seen to correlate with ∼1000 Hz sinusoidal current input.

The impedance sampling schemes varied among the subjects. For S1, impedance was sampled approximately once per day for the first 128 days post-implant and then about once per 15 min for the rest of the recordings. For S2, it was sampled about once per 15 min for the first 3 weeks and then every 5 – 15 min. For S3, one impedance value was measured every 15 min throughout the recordings. S4 did not have impedance measurements for the first 9 days and then it was sampled at about one sample per 15 min. For S5, the impedance was sampled at about every 5 min throughout the recordings. Larger intermittent intervals without impedance measurements existed in the recordings related to the loss of wireless connectivity.

Note that for the analysis of acute and subacute impedance changes, S4 was excluded due to the absence of impedance in the first 9 days post-implant. For the analyses of impedance in gaps of therapeutic stimulation and long-term stability of 24-hour cycles, S5 was excluded because no gap exists in the recordings and S5 does not have adequate data for long-term analysis. Additionally, for the analysis of long-term stability, the first 128-day recording of S1 was excluded due to the low sampling rate, and no analysis of S3 was presented for the left AMG-HPC due to prior resection of the anterior temporal lobectomy.

All activities were approved by Mayo Clinic IRB:18-005483 ‘Human Safety and Feasibility Study of Neurophysiologically Based Brain State Tracking and Modulation in Focal Epilepsy,’ and all subjects provided informed consent.

### Characterization of acute and subacute impedance change

To characterize the initial dynamics of *Z* after implantation, we proposed a piecewise function model consisting of parabolic and exponential functions:

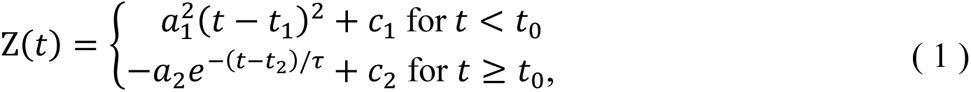

where the model parameters 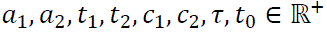. The initial decrease and subsequent increase in effective impedance were modeled by the parabolic function, while the subsequent recovery of impedance until a relatively stable level was described by the exponential function. Node *t*_0_ indicates the time boundary of the two functions. Here, the time constant τ characterizes the rate of the impedance change after *t*_0_. A more intuitive measure is the half-life, *t*_1/2_ = τln(2), which is the time elapsed from *t* = *t*_0_ to the instant when Z arrives at the midway point to the asymptotic value *c*, that is, 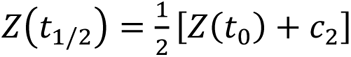. Model parameters were estimated using a nonlinear least-squares algorithm (Appendix A).

By assuming that the impedance reaches the steady state when the first-order derivative of the exponential function is smaller than a threshold α, the start time, *t*_α_ > *t*_0_, of stable Z can be determined as follows (Appendix A):

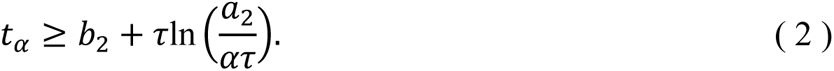

The suggested empirical model is descriptive rather than derived from fundamental physiological mechanisms, and thus may not be unique in characterizing the data. Therefore, we explored an alternative model of double exponentials given by

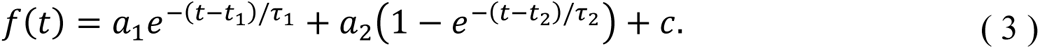

Setting *t*_1_ = *t*_2_ = *c* = 0 results in a four-parameter equation:

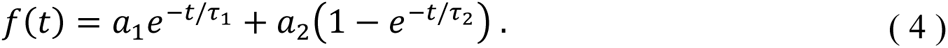

This model is continuous and has less degree of freedom. Figure 2C shows the comparison of the fittings of the two models to the raw impedance data. However, the piecewise model acquires a higher capability of explaining the variance in the data.

**Figure 2.**
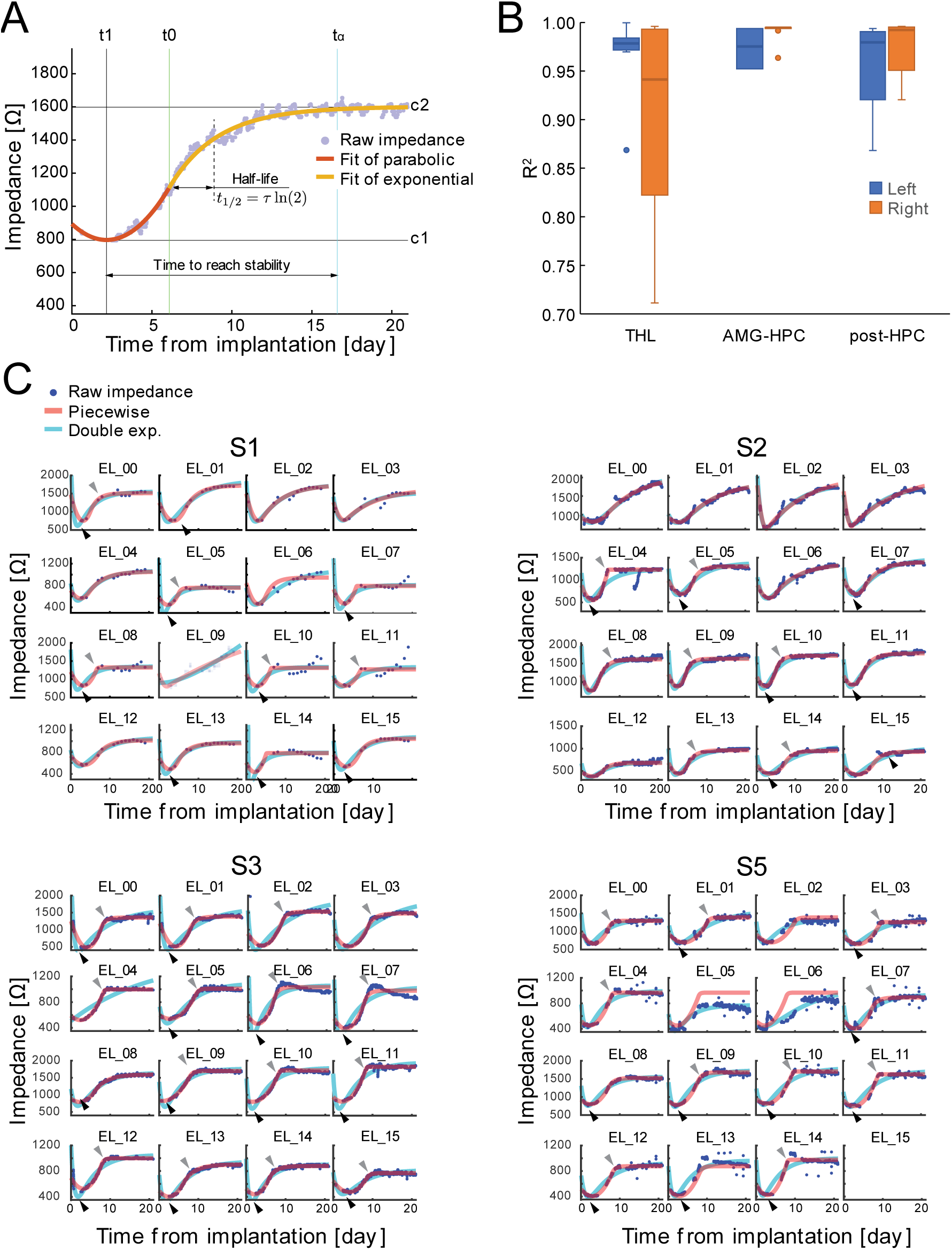
Fitting the model to the impedance change in the first three weeks post-implant (S1, S2, S3 and S5). **(A)** The measured effective impedance values (*Z*) were fitted with a piecewise function consisting of a parabolic and an exponential function, Equation (1). The light purple dots show the sampled impedance measures. The orange curve indicates the fitted parabolic function, while the brown curve the fitted exponential function. *t*_1_is the time when the fitted function is at the minimum *c*_1_, *t*_0_ the boundary between the functions and *t*_αα_ the time when the impedance is supposed to be at stable state (see Methods and Appendix). We define the time to reach stability as the time elapsed from *t*_1_ to *t*_αα_, i.e., *t*_αα_− *t*_1_. *c*_2_is the asymptotic level of the fitted exponential. Note that half-life *t*_1/2_ is relative to the boundary *t*_0_. **(B)** Boxplots of R^2^ (goodness-of-fit, GOF) of the model fits of all individual channels shown in (C) (sample size *N* of THL: [*N*_left_ = 16, *N*_right_ = 16], AMG-HPC: [6, 8] and post-HPC: [10, 7]). **(C)** The raw impedance measures and the fitted models at each individual channel (see Supplementary Table 3 for the locations of the electrodes). The blue dots indicate the raw impedance, the red curves the fitted piecewise model and the cyan curves the fitted double exponential model. Arrows indicate apparent deficiency of the double exponential model. Note that subject S4 was excluded because no impedance measurement was performed in the first 9 days after device implantation and electrode EL_15 of S5 was disconnected after the implant. Abbreviation: **exp.**, exponential.

### Characterizing Z in gaps of therapeutic stimulation

Impedance changes during gaps in therapeutic stimulation provide a means to investigate the properties of electrode-tissues interface and bulk tissue close to the electrode. We define a gap as the time interval when the amplitude of the electrical current stimulation signal is set to zero. The timing and duration of the gaps varied within and across subjects, as the temporary cessation of stimulation was in accordance with each patient’s clinical needs. To select valid gaps for subsequent characterization, we applied two criteria: (1) the gaps were at least 3 weeks (i.e., 21 days) after the implantation of the RC+S™ device, and (2) at least four impedance values were measured in a gap for reliable fitting of an exponential function (described below).

For each valid gap, we quantified the impedance change relative to the impedance before the stimulation was turned *off* and the half-life (*t*_1/2_) of the fitted exponential function. We refer to a unique combination of amplitude, frequency, and pulse width of stimulation as a single state of stimulation. We found the median impedance during the gap and the stimulation state immediately before the gap and calculated the relative impedance change as the difference between these two medians. Additionally, we estimated the half-life of the impedance change by fitting a single exponential (not the piecewise model shown in Section Characterization of acute and subacute impedance change) in a gap. Because the impedance was sampled nonuniformly, the impedance measurements in some gaps were not adequately sampled. To overcome this difficulty, we uniformly resampled the impedance measurements (MATLAB function *resample* with linear interpolation) to 24 samples per day before curve fitting. We used the coefficient of determination (R^2^) as an indicator of goodness-of-fit (GOF), and only included measures of *t*_1/2_from fittings with R^2^ ≥ 0.85 in further analyses.

### Statistics of long-term impedance change in gaps

We adopted the generalized estimating equation (GEE) method, as suggested in a previous study on chronic impedance measurement of a neuromodulation system [7], to estimate the confidence intervals of the time course of long-term impedance variation during stimulation gaps. As an extension of the generalized linear model (GLM), the GEE is suitable for modeling data with high correlation due to repeated measurements and missing data [26]. In our analysis, we followed the paradigm suggested in [7], using binned time intervals as independent variables. The estimation was implemented with the GEEQBox MATLAB toolbox [27] using an identity link, assuming a normal distribution and AR(1) correlation structure. Statistical significance was defined as *p* < 0.01.

### Amplitude and phase of 24-hour circle

To assess the stability of the amplitude and phase of 24-hour circle of effective impedance over time, we partitioned the impedance recording into valid segments. These segments must meet three criteria: (1) at least 3 weeks after device implantation, (2) a minimum duration of 5 days, and (3) at least 40 impedance samples available. The segments were smoothed by resampling the impedance measurements to 24 samples per day (see above for the method). We then applied a Fast Fourier Transform (FFT) to obtain the amplitude and phase of the 24-hour cycle of mean-subtracted impedance in each segment. The estimated amplitudes and phases were grouped every 100 days, with median and boxplot obtained for each 100-day segment. Phase refers to the relative time of the peak of circadian cycle from midnight (12 am), with a phase of 12 pm, for instance, indicating that the peak of the circadian cycle occurs at 12 pm (corresponding to 180°).

## Results

We collected impedance measurements for approximately 1579 days, with a median of 285 days and a range of 124 to 694 days. The average number of impedance samples per day varied between 33.13 and 239.99 (Table 1). Note that during the recording periods, most seizures occurred in the left hemisphere, with the percentage of left seizures being 98.71% for S1, 99.20% for S2, 58.93% for S3, 100% for S4, and 86.58% for S5.

### Significant difference of characteristic measures between left and right hemisphere in initial impedance change

The initial impedance change, which includes acute followed by subacute phase, showed typical biphasic dynamics across all electrodes and subjects. This was characterized by a rapid drop followed by a slower recovery to a stable state, as shown in Figure 2. We fitted the change to a piecewise function, as described by Equation (1) (Figure 2A). We additionally compared it with an alternative model of double exponential, Equation (4), at individual electrode level (Figure 2C). Overall, the piecewise model adequately represented the impedance change (Figure 2B and C; S4 excluded) with an R^2^ = 0.89 ± 0.21 (Mean ± SD, n = 4; see figure caption for sample size at specific locations). After the implant of the device, the impedance reached its the minimum in 2.83 ± 0.63 days (Figure 3A). The time required was shorter in the right hemisphere (2.43 ± 0.39 days) than in the left hemisphere (3.32 ± 0.62 days) of THL (two-sample *t*-test assuming unequal variance: *p* < 0.001, outliers excluded; see Figure 3 caption for sample size), but shorter in the left hemisphere (2.37 ± 0.15 days) of AMG-HPC compared to the right (2.85 ± 0.31 days, *p* < 0.01). No significant difference was found in post-HPC. The half-life measure (Figure 3B) of left THL (2.92 ± 2.91 days) was longer than right THL (0.65 ± 0.66 days, *p* < 0.01). No significant differences were found in AMG-HPC or post-HPC (all *p* > 0.01).

**Figure 3.**
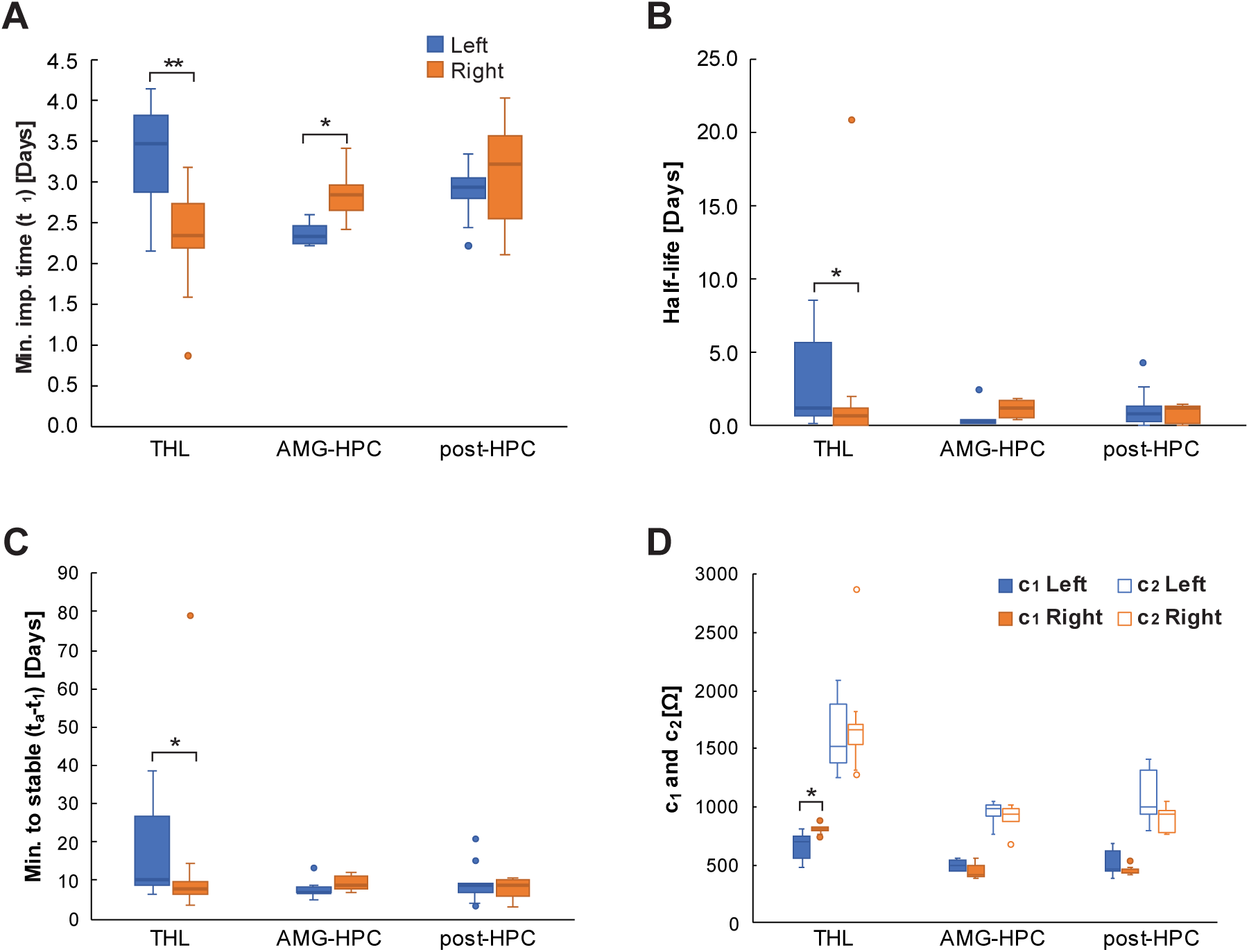
Characterization of acute to subacute impedance change post-implant (S1, S2, S3 and S5). Boxplots of **(A)** time of minimum impedance (*t*_1_) of the fitted models for all the available electrodes from the three areas at left/right hemisphere (sample size *N* of THL: [*N*_left_ = 16, *N*_right_ = 15], AMG-HPC: [6, 8] and post-HPC: [9, 7]), (**B)** half-life ([16, 15], [5, 8], [9, 7]), **(C)** time from the minimum impedance to reach stability ([16, 15], [5, 8], [7, 7]) and **(D)** the minimum (*c*_1_, [16, 14], [6, 8], [10, 6]) and asymptotic (*c*_2_, [16, 14], [6, 7], [10, 7]) impedance levels are shown. The solid-colored boxes are for *c*_1_and the no-filled ones *c*_2_ in (D). For all boxplots, the blue boxes were from channels of the three anatomic areas at the left hemisphere and the orange ones at the right hemisphere. Note that S4 was not included in this analysis (see Methods and Figure 1) and that the sample size *N* involved in significant test excluded outliers (see Supplementary Table 3 for total sample size). Significance test: two-sample *t*-test assuming unequal variances, * *p* < 0.01, ** *p* < 0.001; Abbreviation: THL, thalamus; AMG-HPC, amygdala-hippocampus; post-HPC, posterior hippocampus; Min., Minimum; imp., impedance.

We defined impedance stability as an impedance rate-of-change (calculated as the first-order derivative of the fitted exponential) of less than α = 5 Ω/day. We found that the time to reach stability (*t*_α_− *t*_1_, Figure 3C), where *t*_α_ is given by equation (2), was significantly longer in the left hemisphere (17.17 ± 11.41 days) than in the right hemisphere (8.08 ± 2.88 days, *p* < 0.01). No significant difference was found in the AMG-HPC or post-HPC structures (all *p* > 0.01). we also examined the minimum impedance level (*c*_1_) and the asymptotic level (*c*_2_) (Figure 3D) and found that only *c*_1_ in the left THL (667.58 ± 109.83 Ω) was significantly lower than the right THL (815.63 ± 33.52 Ω, *p* < 0.01). No significant difference was found in the other structures for *c*_1_ and no difference was found in all structures for the asymptotic level *c*_2_.

The statistics related to THL were consistent across subjects and were not biased by individual subjects (as shown in Supplementary Figure 1).

Taken together, these results suggest that, immediately after implantation during the acute and subacute phases, the THL impedance in the more epileptogenic left hemisphere took a longer time to decrease to a lower level than the less epileptogenic right hemisphere. However, it eventually recovered to approximately the same level as the right hemisphere, albeit over a longer period.

### Impedance changes during gaps of therapeutic stimulation

We identified 30, 5, 27, and 29 valid gaps (see Methods) in subjects S1-S4, respectively, as shown in Figure 4 (S5 excluded). Due to the intermittent nature of the temporary pauses in stimulation, the availability of impedance information during the gaps varied among the subjects (Table 2). Figure 4A shows a typical time course of impedance rebound in the gaps close to an implant date. The top row of Figure 3C displays the clusters of impedance changes per subject, as well as the mean and 95% confidence interval (CI) of the impedance change as a function of time, estimated by the GEE model in the stimulation channels. The impedance changes were larger when the gaps were closer to the implant date. The data point of S1 at approximately 600 days indicated no significant changes around that time. Furthermore, no clear correlation was observed between the trend of impedance changes and stimulation conditions immediately before the gaps. This contrasts with the measurements in the non-stimulation channels shown in the lower row of Panel C, where no such impedance change trend was observed. We also conducted a control experiment to assess the effect of the known growth process of a hydrous oxide layer on the iridium surface [28] on the observed impedance difference (Figure 4A). The results suggest that the process was unlikely to be responsible for this observation (see Discussion).

**Figure 4.**
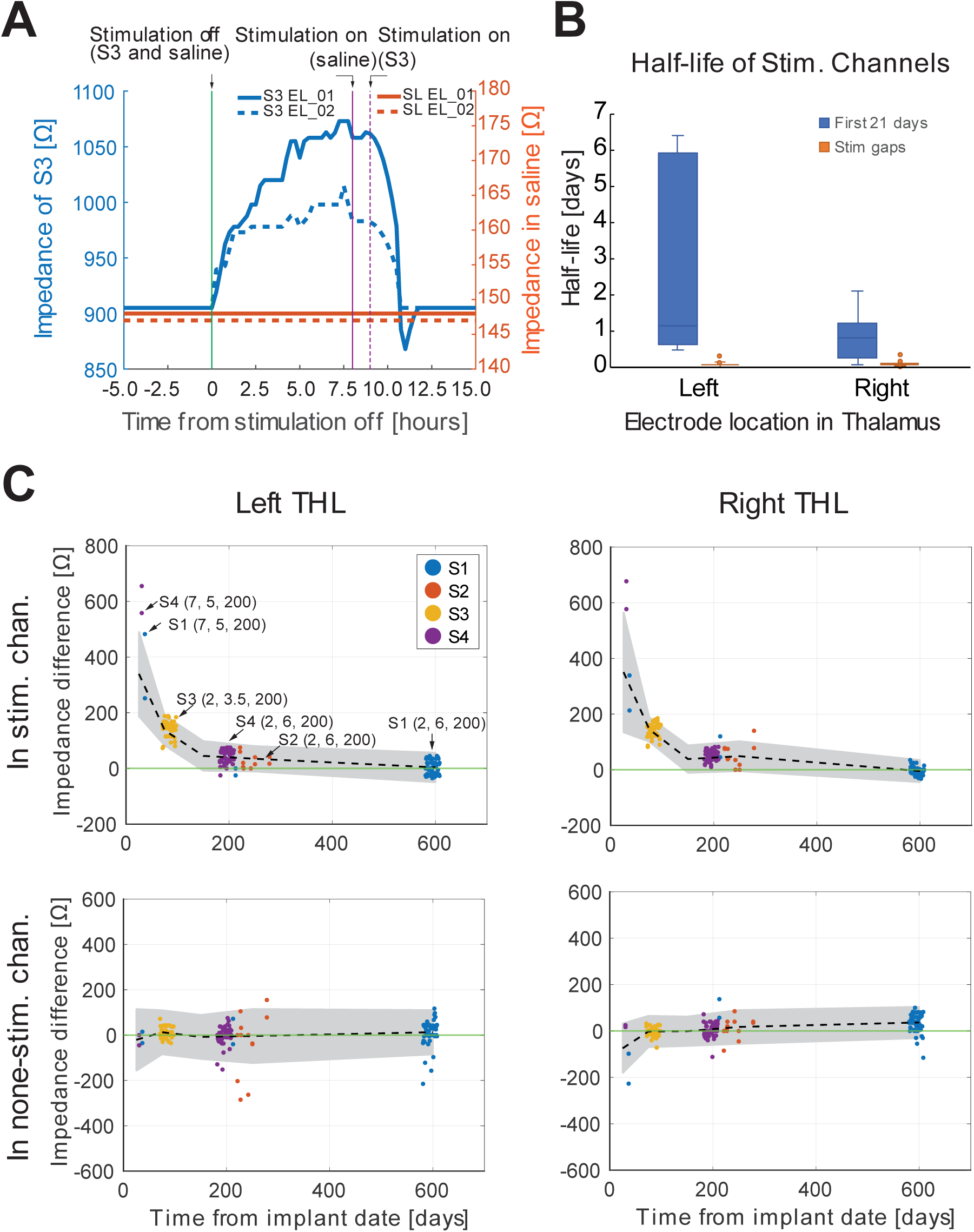
Characterization of impedance change during the gaps of therapeutic stimulation from subjects S1 – S4. (A) An example of impedance changes *in vivo* and in saline. The 2-point impedance was measured every two minutes from the electrodes EL_01 and EL_02 of a Summit RC+S™ device immersed in a body of physiological saline. The red solid and dashed lines represent the measured impedance from these two electrodes (between 145 and 150 Ω, right axis). The vertical solid green line indicates the termination of a stimulation (frequency: 2 Hz, amplitude: 3.5 mA, pulse-width: 200 μsec) delivered for > 5 hours, which was resumed after 8 hours (indicated by the purple vertical solid line). As a comparison, the blue solid and dashed lines represent the impedance values of the two electrodes targeting the left THL of subject S3, aligned with those measured in the saline at stimulation offset (t = 0). The parameters of the stimulation delivered in S3 were the same as those for the saline experiment, except that the therapeutic stimulation resumed after ∼9 hours (the dashed purple vertical line). We can see significant rebound of impedance (with maximum around 1000 to 1060 Ω) in this specific gap (∼70 days post-implant). No such rebound can be seen in the impedance measures in the saline. (B) Impedance changes relative to the impedance values prior to the gaps (sample size of gaps 30, 5, 27, 29; see Methods for details). The upper row shows the impedance changes measured from the stimulation electrodes targeted in left/right THL of the four subjects. The lower row the impedance changes from the non-stimulation electrodes/channels in the THL. Each dot indicates the relative impedance change of median values in a gap. The dashed lines indicate the mean impedance rebound values estimated by the GEE model as a function of time, where the shaded areas indicate the 95% CI around the mean. A tuple of parenthesized three values of frequency (Hz), amplitude (mA) and pulse-width (µsec) display the stimulation states immediately before the gaps. For instance, a tuple of (2, 3.5, 200) indicates 2 Hz current pulse with 3.5 mA and 200 µsec pulse-width. (C) Boxplots of half-life measures from the stimulation electrodes. The blue plots show the half-life measures of the fitted exponential functions for the impedance change in the first 3 weeks (21 days) after the implant and the orange plots the half-life measures of the fitted exponential for the impedance in the gaps (sample size *N* of left: [first 21 days = 7, gaps = 108], *N* of right: [7, 86]). Note that all measures are from the stimulation electrodes targeted in THL and that subject S5 was not included in the analysis (see Methods). Abbreviations: THL, thalamus; stim., stimulation, chan., channels; CI, confidence interval.

**Table 2.** Impedance changes during the gaps of therapeutic stimulation.

| Gap cluster |  | Channel | S1 | S2 | S3 | S4 |
| --- | --- | --- | --- | --- | --- | --- |
| 1 | Time [day] |  | 61.00 | — | 84.29 | 30.45 |
| | Left imp. [$\Omega$] | Stim. | 186.25 ± 163.07 | — | 136.59 ± 39.65 | 606.50 ± 68.59 |
| | | Non-Stim. | -7.33 ± 26.14 | — | 8.77 ± 22.40 | -22.50 ± 31.82 |
| | Right imp. [$\Omega$] | Stim. | 192.08 ± 87.39 | — | 140.37 ± 38.00 | 627.00 ± 70.71 |
| | | Non-Stim. | -53.92 ± 97.07 | — | -3.77 ± 17.62 | 21.00 ± 5.66 |
| 2 | Time [day] |  | 212.47 | 241.08 | — | 194.91 |
| | Left imp. [$\Omega$] | Stim. | -12.50 ± 17.68 | 22.83±25.71 | — | 45.94 ± 21.76 |
| | | Non-Stim. | 16.00 ± 79.20 | -38.17 ±141.18 | — | -0.64 ± 37.54 |
| | Right imp. [$\Omega$] | Stim. | 82.50 ± 53.03 | 51.58 ± 39.82 | — | 51.18 ± 18.87 |
| | | Non-Stim. | 97.00 ± 56.57 | 11.67 ± 44.07 | — | 3.99 ± 26.16 |
| 3 | Time [day] |  | 594.47 | — | — | — |
| | Left imp. [$\Omega$] | Stim. | 3.13 ± 25.21 | — | — | — |
| | | Non-Stim. | 14.91 ± 56.50 | — | — | — |
| | Right imp. [$\Omega$] | Stim. | -3.66 ± 13.51 | — | — | — |
| | | Non-Stim. | 37.04 ± 39.62 | — | — | — |

Note: mean ± SD. Abbreviations: **imp.**, impedance; **stim.**, stimulation.

We further compared the estimated half-life values in the stimulation channels between those estimated during the first three weeks post-implant (Figure 3B) and those in the stimulation gaps (Figure 4B). It is important to note that neither therapeutic stimulation was applied in the first 3-week period nor during the gaps, although stimulation was delivered before the gaps. In the right hemisphere, which is the less epileptogenic brain region, the first 3-week half-life estimates were significantly longer than those during the gaps (Right:1^st^ 3-week Mean ± SD, 22.72 ± 15.25 hours; gaps 1.92 ± 1.06 hours; two-sample *t*-test, *p* = 0.01). A similar trend was observed in the left hemisphere electrodes, but with larger variance (Left:1st 3-week 71.78 ± 68.69 hours; gaps 1.76 ± 0.94 hours; *p* = 0.04), suggesting that different mechanisms responsible for the increase in impedance.

### Amplitude and phase of 24-hour cycle appear to be stable over extended periods

We analyzed the amplitude and phase of the long-term 24-hour cycle of impedance, as shown in Figure 5 (also Supplementary Figure 3; see Methods for subjects involved in this analysis). Figure 5A displays the amplitude over the observed recording periods on both sides of THL, AMG-HPC, and post-HPC. The amplitude varied across the subjects, with S1 having a significantly higher amplitude than S2, S3, and S4 in all areas except for the left THL. However, for each individual subject, the amplitude appeared to remain stable throughout the entire observation period. No significant difference was found within the subjects (Wilcoxon rank sum test: all *p* > 0.01, Bonferroni corrected).

**Figure 5.**
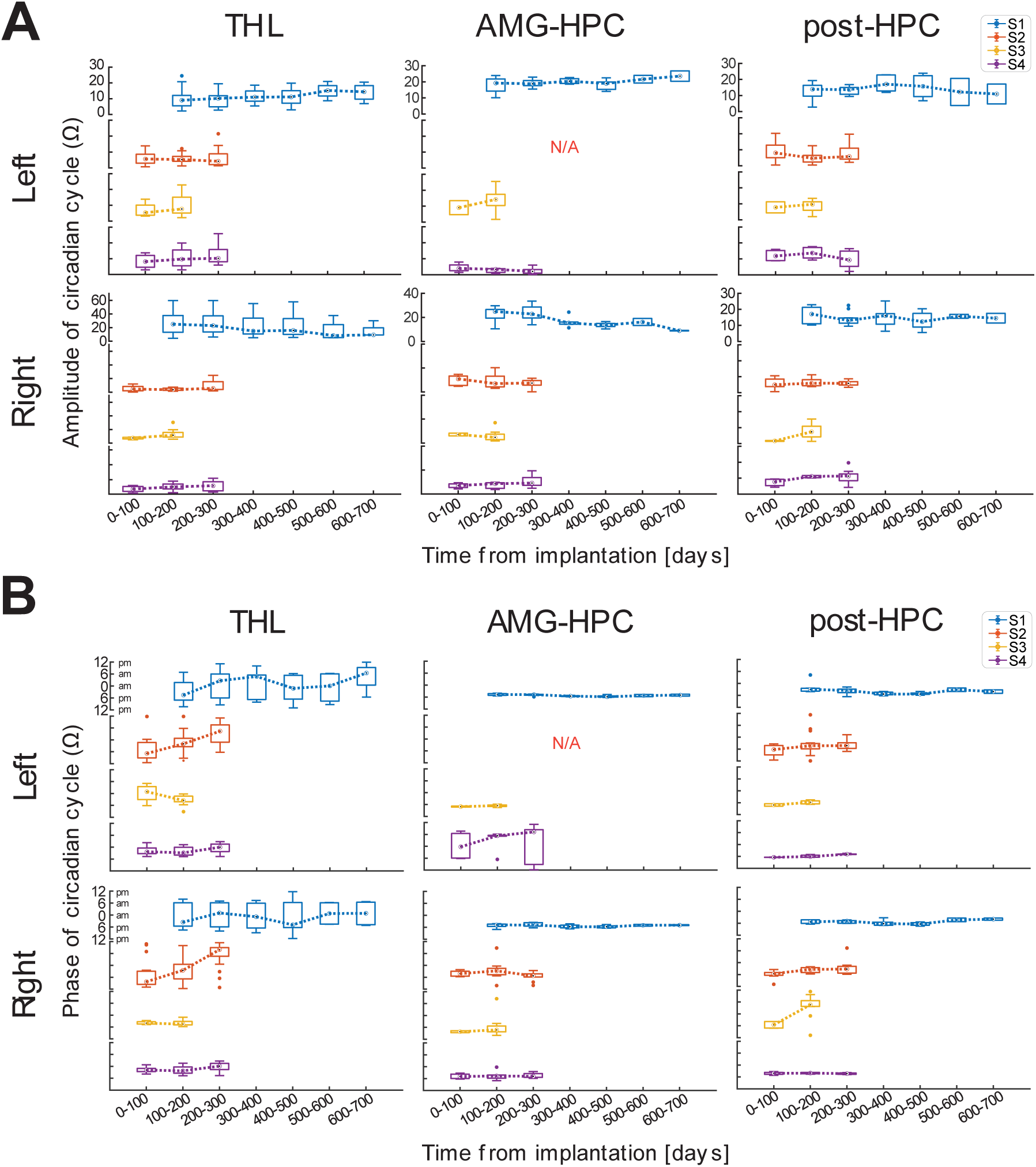
Long-term amplitude and phase of circadian cycles of impedance at THL, AMG-HPC and post-HPC. (**A**) Amplitude and (**B**) phase of circadian cycle of left/right hemisphere. Boxplots represent the distribution of the estimates (see Supplementary Table 4 for the number of samples) in a 100-day interval for each subject. Note that scales vary between panels for amplitude and are consistent for phase. S5 was excluded and no signals from left AMG-HPC structure of S2 (see Methods). Abbreviations: SD, standard deviation; THL, thalamus; AMG-HPC, amygdala-hippocampus; post-HPC, posterior hippocampus.

Regarding the phase values shown in Figure 5B, we observed a similar pattern to that of amplitude. The phase of the circadian cycle remained stable over the observation period, although it varied across different anatomic locations. At THL, the phase values of all subjects did not significantly deviate from 12 am, indicating that the maximum impedance of the 24-hour cycle occurred around midnight. However, the time of maximum impedance differed among the hippocampal structures. Except for S4 left AMG-HPC, which was approximately 12 am and significantly different from S1 and S2, the phase values of S1 and S3 in the left AMG-HPC and of all subjects in other hippocampal structures did not significantly deviate from 6 pm (Wilcoxon rank sum test: all *p* > 0.01, Bonferroni corrected), indicating that their maximum amplitudes were around 6 pm, about 6 hours ahead of those in the thalamus. Supplementary Figure 3 further illustrates the relationship between measured impedance and sleep/awake states. The two-day impedance measurements display an approximate 24-hour cycle, with most Awake states occurring when impedance was higher and most Sleep states occurring when impedance was lower. This is consistent with the reports using animals [18].

## Discussion

In this study, we sought to characterize impedance changes in both short-term (in the first 3 weeks post-implant) and long-term (124 – 694 days) periods in human subjects. We developed a piecewise function to describe the biphasic dynamics of the initial acute to subacute impedance response. Our results suggest that the left and right THL exhibited distinct impedance change dynamics immediately after implantation. The analyses of impedance behavior during the temporal pauses of stimulation indicated an interaction between the stimulation and the tissues near the electrodes and suggested that tissue encapsulation around the electrodes matured at about 200-300 days post-implant. Finally, we found that the amplitude and phase of the prominent 24-hour cycle of impedance were relatively stable in the long term, which is also important for sleep behavior [8, 23] (see examples in Supplementary Figure 3).

### The model revealed differential dynamics of acute and subacute impedance responses in the left and right hemispheres

The short-term change in impedance after lead implantation, characterized by a rapid decrease followed by a slow increase, has been reported in previous studies [6, 7, 13]. The initial drop is thought to result from an inflammatory response to injury trauma caused by the implant [13]. During the early phase of the body’s inflammatory response to foreign materials, an increase in vascular permeability leads to fluid accumulation [29], which in turn reduces the impedance around the electrodes. However, the slow increase in impedance was likely due to the formation of encapsulation layers near the electrodes. The specific morphology of the encapsulated tissue depends on the surface texture, shape, and material of the implant. Changes in the chronic implant tissue interface have been attributed to the growth of the fibrous tissue capsule [13, 30]. While the pattern of the subacute response is well documented, we are not aware of other studies that have investigated long-term properties with sufficient temporal impedance sampling to resolve the more subtle changes that were revealed by the model (discussed below).

By fitting our piecewise model (see Methods), we were able to gain further insights into the response and to provide possible means to compare the results from different studies. For example, the stability of the impedance recovery is not well defined but can be suggested by setting a threshold (in this case, 5 Ω/day) for the first-order derivative of the exponential function. Our modeling results show that using Pt-Ir electrodes, the impedance reached a minimum value at approximately 3 days post-implant (Figure 3A) in human subjects, which is close to 4 days of resistivity measurement using silicon rubber and 2 days using epoxy arrays in cats [13]. The impedance measurements during the first week after implantation in our data were consistently lower than the stable values after 3 weeks, consistent with a previous report of human data from NeuroPace RNS™ device, which showed impedance values in the first week were significantly lower than the stabilized value at one-year post-implant [7]. These results may indicate that similar time courses of adaptation correlated with the progressive development of fibrous tissue capsule around the implants, leading to an increase in impedance/resistivity. This process may not be strongly related to the species or electrode materials. Our data also show that the recovered impedance could reach a stable level in approximately 3 weeks (Figure 3C), which is similar to previous reports of approximately 3 [22] to 4 weeks [31]in rats, approximately 40 days in cats [13], and approximately 4 weeks in humans [7].

However, previous studies [6, 7, 30] did not differentiate their measurements in relation to the degree of epileptogenicity of the tissue and networks involved in the generation of seizures (Supplementary Table 1). Our results indicate that, on average, the impedance in the THL in the more epileptogenic brain network (left side) took longer to reach the minimum than the contralateral, less epileptogenic side of the THL (right side), and a longer half-life and longer period from the minimum to the stable level in the left than in the right side of the THL. No significant differences were observed in the AMG-HPC and post-HPC areas (Figure 3A, B and C). In the THL, the minimum impedance on the left side was significantly lower than that on the right side, but not in the AMG-HPC and post-HPC areas (c_1_ in Figure 3D). No significant difference was observed at a stable level (c_2_ in Figure 3D) in any area. It appears that the observed longer time needed to reach the minimum in the more epileptogenic left THL is mainly due to the lower minimum impedance level, as no significant difference in rate of impedance decrease was found between the left and right hemispheres (see Supplementary Figure 2).

The reason for the difference in acute-to-subacute impedance changes between the left and right THL cannot be directly determined with these data. However, for our subjects, the left hemisphere limbic networks were more highly epileptogenic, with significantly more interictal epileptiform activity, seizures, and more severe delta frequency (Table 1 and Supplementary Table 1). Interestingly, previous studies have argued that the cell types and structure of the encapsulated tissue are largely independent of the site of electrode implantation [13]. If this is true, we speculate that the difference observed during the first 3 weeks post-implant may be related to the pathology of brain tissue in epilepsy, which is known to exhibit significant immunologic dysregulation [32–35]. Impedance changes immediately around seizure spread have been well described [19, 36, 37], but little is known about the interictal impedance characteristics of the epileptic brain. Our findings provide additional evidence and insights into this area of research, which warrants further investigation.

### Analyses of gap impedance indicate maturity of encapsulation layers about 200-300 days post-implant

Intermittent gaps in therapeutic stimulation were present in the recordings for clinical purposes. These gaps provided time windows for checking the impedance characteristics around and within the gaps in electrical stimulation. Our results show that significant rebounds of impedance, from lower values when electrical stimulation is active to progressively higher values after stimulation is turned off, can be observed within approximately 300 days after implantation in the THL electrodes used for stimulation (Figure 4C upper row). As shown in Figure 4A, the control experiment suggested potential interactions between the stimulation and the electrode-tissue interface. The observation likely reflects the previously reported phenomena where voltage applied to the microelectrodes reduces the impedance by “cleaning” the electrode of biological material. The impedance may be further effected by elevated blood flow, as neuronal hyperactivity due to stimulation may increase blood perfusion around the electrodes [38]. Therefore, impedance recovery during the stimulation gaps may be facilitated by decreased metabolic demands and blood perfusion in brain tissue.

Our results suggest that, at least for clinical macroelectrodes, the growth of fibrous capsulation tissue around the electrodes is less dynamic after ∼300 days. The phenomenon of decreasing impedance with voltage biasing (i.e., passing currents) may no longer be viable after ∼1 year. For functionally encapsulated microelectrode sites, it is possible to increase transient conductivity pathways through the encapsulation of tissues by applying a DC bias voltage (typically +1.5 V) to the electrode site for several seconds, known as the “rejuvenation” approach [24, 39]. However, we speculate that the window for this phenomenon for the microelectrodes used to record single neurons [24] is likely earlier and may not be viable for chronic implants.

The therapeutic neurostimulation used in our study was delivered as a counter-balanced square wave pulse (typically with a frequency of 2 Hz, amplitude of 3.5 mA, and pulse-width of 200 µs) for extended period (days to weeks). It is not clear to what extent the stimulation might be able to create transient conductivity pathways that lower impedance. However, given that no significant rebound was found in non-stimulation channels in THL (Figure 4C, lower row), AMG-HPC, or post-HPC (data not shown), it is likely that therapeutic stimulation was able to increase conductivity pathways through encapsulation, particularly in the early period after implantation. After 200-300 days of implantation, with the continuous growth of encapsulated tissue, the creation of conductive pathways could not be established, and the efficacy of tissue stimulation largely diminished. As a result, no significant impedance rebound was observed during the gaps, indicating the maturity of fibrous encapsulation.

Further analysis revealed that the estimated half-life values in the gaps (1-2 hours) were significantly shorter than those estimated in the first 3 weeks (several days, Figure 4B). These results suggest that different mechanisms and stages of the encapsulation processes may be responsible for the increase in impedance. Our findings may be useful in guiding the delivery of therapeutic stimulation at various times post-implant.

### Long-term stability of 24-hour cycle of impedance

One challenge of chronically implanted neuromodulation devices is the progressive development of encapsulation layers around the electrodes, which can sometimes electrically shield an electrode from the surrounding tissues, affecting the LFP sensing and therapeutic efficacy of neuromodulation. This condition may be monitored by periodic impedance measurement, and some previous studies with very sparse sampling of impedance have reported that impedance is largely stable over long-term follow-up [6, 7]. In this study, we densely sampled impedance and examined the stability of long-term impedance from a novel perspective by studying the stability of the amplitude and phase of the circadian cycle of brain impedance.

It has been recently shown that 24-hour cycle of the effective impedance is a prominent feature with little variation in periodicity over the duration of recording [23]. However, variations in the amplitude and phase of the circadian impedance cycle have not been fully investigated. This is important because the effect of the changes of bulk tissue (brain matter and encapsulation tissue) near the electrodes may not have a major influence on the periodicity of the cycle, but rather on the properties of electrical impedance, which are reflected by the variation in the amplitude and phase. Our results showed that the amplitude and phase were stable within the subject over the long-term recording period. Since the circadian cycle is thought to be related to variations in extracellular space volume associated with the sleep-wake cycle [21, 23], the stability of amplitude and phase suggests that the impedance variation due to the growth of encapsulation layers in the immediate vicinity of the electrodes was substantially smaller than that due to the circadian cycle. Therefore, when evaluating controlled stimulation design or tissue-electrode interface properties by analyzing impedance measurements, it is recommended to preprocess the data by removing the circadian component.

### Limitations of this study

A major limitation of our study is the technical challenge of clearly differentiating between the impedance of the electrode-tissue (i.e., electrode-electrolyte) interface and that of bulk tissue, which includes brain matter and encapsulation layers (if formed), due to the use of two-point impedance measurement. We recently investigated the differences between 4-point and two-point method in saline and saline-microbead composite materials (Figure 1; also see Methods in [23]). Our results show that the measured impedance with Medtronic Summit RC+S™ is insensitive to the impedance of electrode-tissue interface. Therefore, we attribute the observed impedance dynamics mainly to changes in bulk tissue near the electrodes. Furthermore, our observed half-life values of acute-to-subacute impedance change after implantation (Figure 3), impedance change during stimulation gaps (Figure 4), and relatively stable circadian impedance cycle (Figure 5) indicate that changes in electrode-tissue interface impedance are unlikely to be the main cause of the observed dynamics.

There are several other limitations to this study that should be noted. First, all the patients in our study had drug-resistant mesial temporal lobe epilepsy, and our results may reflect the response of the epileptogenic brain and may not be applicable to other neurological and psychological disorders treated with electrical brain stimulation (EBS). It is notable that there was a difference between the left and right hemispheres, which may reflect the greater epileptogenicity (tendency of seizures and interictal epileptiform discharges, IED) of the left hemisphere in these subjects. Second, the etiologies and functional and structural imaging findings of the patients were heterogeneous, which may impact impedance. Additionally, they were taking different anti-seizure medications that may affect brain impedance. Where possible, medications remained fixed in this study. Third, the targeting of the electrodes is accurate to approximately 2-3 mm. The impact of the imaging resolution of brain substructures and nuclei on impedance measurements cannot be ascertained with the current data. Finally, we did not directly investigate the impact of seizures and IED on impedance, which is an area of current investigation.

## Conclusion

Characterizing the full dynamics of impedance is important for understanding its impact on LFP sensing, therapeutic electrical stimulation, and for ensuring efficient and stable electrical brain stimulation. In this study, we densely sampled the impedance and developed novel approaches for analyzing the time course of the impedance response from the acute (1-3 days) to subacute (4–3 weeks) and long-term (> 3 weeks) stages.

For short-term (acute and subacute) changes in impedance, our results largely support the previous findings. We further characterize the impedance response by fitting a piecewise function, where a parabolic captures the drop and initial rebound phase and an exponential to approximate the later asymptotic phase of the impedance. This descriptive model may be useful for comparing the results from different experiments. It is worth noting that, according to the estimates from the model, significant differences in characteristics between the left and right THL are present. We speculate that this reflects the more epileptogenic left hemisphere limbic network and AMG-HPC structure. Given the dominant left seizures in our subjects, these findings may have implications for the pathological effect on short-term impedance dynamics, which requires further investigation.

In our investigation of long-term impedance measures, we found significant impedance rebound during the temporary gaps of the stimulus, consistent with other studies. However, our results further suggested that the degree of rebound decreased over time and was no longer observable between 200 and 300 days after implantation, indicating possible maturity of encapsulation by fibrous tissues around the electrodes. We also propose a novel perspective on long-term impedance by examining the stability of the amplitude and phase of the prominent circadian cycle of impedance. Our findings suggest not only that the amplitude and phase were relatively stable over time, but also that the daily variance was dominant in impedance changes.

## Data Availability

Data are available upon reasonable request to the authors.

## Acknowledgments

We thank Irena Balzekas, Victoria S. Marks PhD, Jordan S. Clark MS, Andrea M. Duque Lopez MD, and Dalia Zubidat MD for their thoughtful discussions during this study and preparation of the manuscript.

## Data accessibility

Data are available upon reasonable request to the authors.

## Appendix

### A. Estimate model parameters and time of stability of subacute impedance change

We estimated the model parameters of Equation (1) using the nonlinear least-squares algorithm (MATLAB^©^ Function *fit*). To fit the model, we assume that *Z*(*t*) and its first-order derivative *Z*^′^(*t*) are continuous at node *t*_0_, the boundary of the parabolic function, and the exponential function, that is,

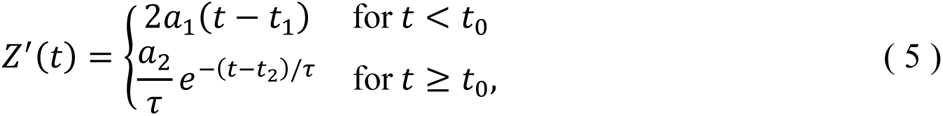

and

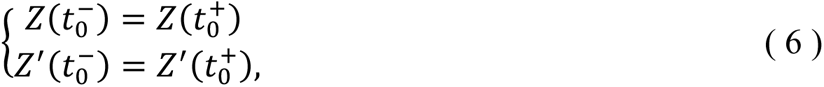

which gives us the relations of *t*_0_ and *a*_2_ with other parameters,

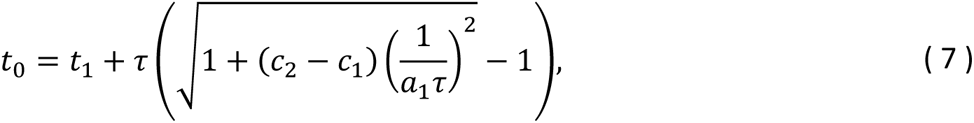

and

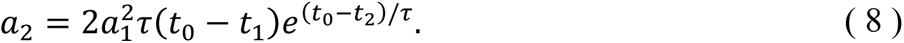

Thus, six parameters, 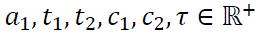 were estimated from the model fitting. The initial values of these parameters are presented in Supplementary Table 2.

From Equation (3), if we define that *Z*(*t*) is stable when its change rate is below a threshold α, i.e., *Z*^′^(*t*_α_) ≤ α for *t* ≥ *t*_0_, then we can find the start time of stable *Z*(*t*) shown in Equation (2). In practice, α was arbitrarily set to 5 Ω/day.

## Footnotes

1 In this paper, the term “24-hour cycle” is used interchangeably with “circadian cycle”.

